# Symptoms network analysis of serious mental illness: A cross disasters comparison

**DOI:** 10.1101/2021.04.10.21255242

**Authors:** Yafit Levin, Bachem Rahel, Robin Goodwin, Menachem Ben-Ezra

## Abstract

**Background:** The Kessler Psychological Distress Scale (K-6) has been used worldwide in community epidemiological surveys and served as a screening measure for serious mental illness in the general population. We take a novel approach by examining the symptoms network of the K-6 and the exploration of differences between three types of disasters: Nature related, Terror attacks, and COVID-19.

**Aims:** To explore the K-6 symptoms network and its structure replication across the three types of disasters.

**Methods:** A network analysis of psychological distress symptoms as assessed by the K-6 was conducted using data from 9,271 participants from different disaster samples: Terror (n = 5842), COVID-19 (n = 2428), and Nature related (n = 1001).

**Results:** While there were extensive connections between items across all disaster samples, network structure differed across the disaster types. While after a nature related disaster and the COVID-19 pandemic depression- and anxiety-items were interconnected, a terror attack resulted in more separated manifestations of anxiety and depression. Centrality analysis showed “depressed/no cheering up” to be the node with the highest strength centrality in all networks; in the Nature-related network, “restless or fidgety” was also highly central.

**Conclusions:** Results provide evidence of different psychological distress structures in different disasters. Depending on the type of disaster, trauma-focused interventions may need to be augmented, with specific components directed at depression and/or anxiety.

## Introduction

Communities all over the world endure all-encompassing aversive and traumatic events, including global pandemics, disasters following natural events, and terror attacks. All of these can trigger significant psychological distress, with disasters following natural events the potential to quickly overwhelm healthcare resources ^1^. The current study explored differential stress reactions, as assessed by the 6□item Kessler Psychological Distress Scale (K-6) ^2^, to three different types of disasters: disasters resulting from natural events (flood), terror attacks and distress associated with pandemic threat (COVID-19). This work takes a novel approach by examining the symptoms structure of the K-6 and explores connectedness between symptoms and centrality across three types of disasters. This will allow practitioners to rapidly identify core symptoms of interest, by exposure event, and thus more target interventions.

### The Kessler Psychological Distress Scale

The Kessler Psychological Distress Scale (K6) is a dimensional measure of nonspecific psychological distress used extensively in epidemiological surveys of adults in the community ^2^. Widely used by the WHO for screening for serious mental illness ^3^, the scale outperforms other screening measures (e.g., General Health Questionnaire) in predicting severe mental illness, with high levels of sensitivity and specificity ^4,5^. However, some ambiguity exists regarding the factor structure of the K6. While factor models and reliability analyses mainly confirmed a one-factor structure ^6,7^, other analyses support a two-factor structure, with four items representing depression and two items representing anxiety ^8–10^. This has led to the establishment of preliminary cutoff scores for the K6-depression and K6-anxiety subscales ^10^. Thus, the K6 appears to have clinical utility by using a total score, subscale scores, or both.

In recent years, a novel technique emerged to assess the structure of mental health questionnaires beyond factorial latent analysis: Symptoms Network Analysis. Network analysis allows for the analysis of the complex interactions between individual symptoms that characterize psychopathology as opposed to methods that rely on the identification of the latent structure of psychopathology ^11^, and the assessment of the role of specific symptoms in activating other symptoms. This approach may shed further light on the symptoms structure of psychological distress as assessed by the K-6. Network analysis not only allows exploring the symptoms network structure of the K-6 but also allows us to identify the most central distress symptom for each disaster type (i.e., the symptom that is most strongly related to other symptoms in the network). A network analysis of the K-6 can highlight possible factors of depression, anxiety, and psychological distress, by identifying highly connected items vs weaker connections, as well as connecting or “bridging” items. Disasters often present demanding situations which overload services, requiring rapid identification of necessary interventions. Knowing the importance of anxiety rather than depression or a blend of the two which represents psychological distress, during a specific type of event is therefore important for intervention strategies, as well as making predictions about processes of recovery and prognosis ^12,13^.

### Types of disasters and mental health impact

According to the World Health Organization, a disaster is defined as a serious disruption of the functioning of a community or a society causing widespread human, material, economic or environmental losses which exceed the ability of the affected community or society to cope using its own resources ^14^. Disasters are social and environmental processes often subdivided into natural processes or human-made events ^15^.

Nature related disasters are increasing in frequency and impact, causing substantial disruption in many countries ^16^. In the last decade, more than 2.6 billion people have been affected by natural phenomena such as earthquakes, tsunamis, landslides, cyclones, heat waves, floods, or severely cold weather. It is increasingly recognized that nature related disasters have a human-made component that reflects inequities in infrastructure, environmental risk-taking, local politics and culture ^17,18^. Systematic reviews and meta-analyses have found that both general psychological distress and the prevalence of psychiatric disorders increased after nature related disasters ^19,20^.

Relying on the above definition of disaster ^14^, a pandemic can be viewed as both natural ^21^ but also human-made. Although events such as a hurricane may precipitate a long chain of cascading sequela ^22^ and infectious disease ^23^, COVID-19 is particularly distinctive for the considerable duration of the continued threat, accompanied by a series of non-pharmaceutical interventions, such as self-isolation, quarantine and social distancing, that, alongside the threat of mortality and morbidity, can cause additional psychological distress ^24–26^. COVID-19 is also distinctive because of the particularly high levels of uncertainty over both the levels of threat and the appropriate response ^27^.

Terror attacks, however, are intentional human-made events that can be distinguished from natural occurrences such as earthquakes or a novel zoonosis. Terrorism creates complex scenarios of harm that involve both “targeted hatred and random violence” ^28^. Particularly notable, terror attacks can change perceptions of other people and their intentions, thus raising fundamental moral questions. By undermining world assumptions ^29,30^, and suggesting we live in a world that is malevolent and unexpected ^31^, mass terror events can overwhelm defense mechanisms and also lead to psychological distress ^30^. Alongside these unique features, terror events can also of course post challenges similar to those posed by other types of large-scale catastrophes, including the financial and operational challenges involved in repairing infrastructural damage ^32^, and causing reputational harm to a location or population.

Though studies typically measure the degree of distress after disasters, it has also been argued that the form of psychological distress may also be significant for understanding the phenomenon ^33^. To date we know of no previous study that has compared different types of trauma with regards to varying forms of distress. Whereas studies that have endeavored to explore differential forms of distress have for the most part focused on differentiating a depressive form of distress and a more anxious form ^34^, a data-driven examination using network analysis may reveal subtle but critical clinical interactions between symptoms.

### Aims of the study

In this study we examined the following research questions: (1) what is the pattern of interactions between the K-6 psychological distress symptoms - Are depression, anxiety or general psychological distress (i.e., a blend of anxiety and depression) most closely associated with a particular type of disaster; (2) do the symptoms network interactions replicate across different types of disasters; (3) which symptom is the most central and how does this replicate or differ across networks. Answering these questions will facilitate informed judgment about the components most relevant for scalable interventions in the aftermath of diverse types of disasters (i.e., provide information about which symptoms should be targeted in the aftermath of a specific disaster type).

## Methods

### Participants

#### Nature related disaster sample from the Philippines

An online panel survey was conducted in the Philippines three weeks following the Super Typhoon Haiyan (beginning on November 27, 2013) in collaboration with Asia Opinions, a survey company specializing in East Asia. Inclusion criteria were age 18 or older and living in areas affected by typhoon Haiyan. The panel was created using random stratified sampling methods, using weights for key demographic elements (e.g., gender, region) that were compared with census information to create a reliable approximation of a representative sample. The procedures followed those established by the ICC/ ESOMAR International Code on Market and Social Research ^35^. Of a total of 1,400 individuals contacted, 1,001 completed the survey. More information on demographics can be found elsewhere ^36^.

#### COVID-19 samples from China and the UK

Following ethical approval by the funding university, two online surveys were conducted, with a random and stratified sampling, between March 30 and April 2^nd^ 2020, one week after national lockdowns: China (N=1135) and the UK (N=1293). In both countries, the same survey company (AsiaOpinions) accessed a series of internet panels in each country, following guidelines established by the International Chamber of Commerce International Code on Market and Survey Research (ICC/ESOMAR). Online consent was obtained for all participants. All procedures complied with the Declaration of Helsinki 1975/2008. The mean age of the participants was 51.51 years (SD = 14.75, range 18–75), 53.3% were female (n = 689), 69.0% reported being in a committed relationship (n = 892) and 27.2% (n = 352) reported having a background medical condition (hypertension, diabetes, cardiovascular disease, chronic respiratory disease, chronic obstructive pulmonary disease and cancer). Demographic description of the two data sets can be found in ^37^.

#### Terror samples from France, the UK, and the US

Here the sample comprised of a series of data collections following a) two terror attacks in France during 2015, b) a terror attack in the UK in May 2017, and c) a terror attack in the US on June 2016. A combined sample size of 5842 participants represented general populations in each country following terror attacks. Further details of each sample are provided in Online Supplementary Materials and elsewhere for the samples of the UK ^38^, the US ^38^ and France ^39^.

### Measures

#### Six□item Kessler Psychological Distress Scale

The 6□item Kessler Psychological Distress Scale (K□6) has been validated to assess nonspecific psychological distress during the past month, including symptoms of anxiety and depression ^2,5^, and used to screen for psychological distress ^7,40^. The scale includes 6 psychological symptoms: “nervous”, “hopeless”, “restless or fidgety”, “so depressed that nothing could cheer you up”, “everything is an effort” and “worthless”. participants respond on a 5□point Likert scale (“0” = never, “1” = occasionally, “2” = part of time, “3” = most of time, and “4” = all the time), with a total score ranging between 0□to□24 ^5^. A total score > 12 is defined as severe psychological distress ^41^. The has been found to be reliable with Cronbach’s a values of .89–.92 ^2^. The current samples revealed strong reliability with Cronbach’s alphas of .90 to .92 in all samples.

### Statistical analysis

#### Regularized partial correlation networks across the total sample and the three disaster type samples

##### Network Estimation

The symptom networks were estimated for all symptoms of the K-6, for the entire sample and then for the three subsamples which adhere to different types of disasters (Nature related disaster, COVID-19 pandemic, and Terror) using the R-package qgraph ^42^. Networks were estimated using regularized partial correlation models in the R-package qgraph that present the unique, independent relationships between symptoms ^43^. Symptoms in a network model are nodes; associations between nodes edges. Due to the cross-sectional nature of the study we use an uweighted and undirected network. Questionnaire data were answered on an ordinal scale; thus, we estimated a polychoric matrix. We estimated partial pairwise correlations parameters between all nodes through a Gaussian Graphical Model (GGM) ^43^.

##### Visualization with the Graphical Lasso

We used the graphical least absolute shrinkage and selection operator (glasso; implemented in qgraph), which visualizes sparse networks using part correlations and allowed for the ordinal scale of the questionnaire. This method directly estimates the inverse of the covariance matrix ^44^, shrinking small edges and key parameters to zero by estimating a penalized maximum likelihood solution based on the Extended Bayesian Information Criterion (EBIC) ^45^. For ease of visual comparison, networks were restricted to a consistent “average layout,” presented across all samples.

##### Network stability

We examined the stability of the individually estimated networks, including estimating 95% confidence intervals around the edge weights and estimating a correlation-stability coefficient for strength centrality.

##### Network Comparisons

To compare differences between networks, we estimated network differences between each pair of networks using the *NetworkComparisonTest* (NCT) package in R ^46^.

## Results

### Regularized partial correlation networks among the total sample and across the three types of disasters

#### Network estimation

To enhance visual comparability of edges, we estimated the average layout of the three networks and presented all networks using this layout (see Fig. 1). In all four networks the K-6 symptoms network showed that 13-15 of 15 possible edges were nonzero (86%-100%). This suggests the symptoms had extensive connections with each other in all samples.

**Fig 1.**
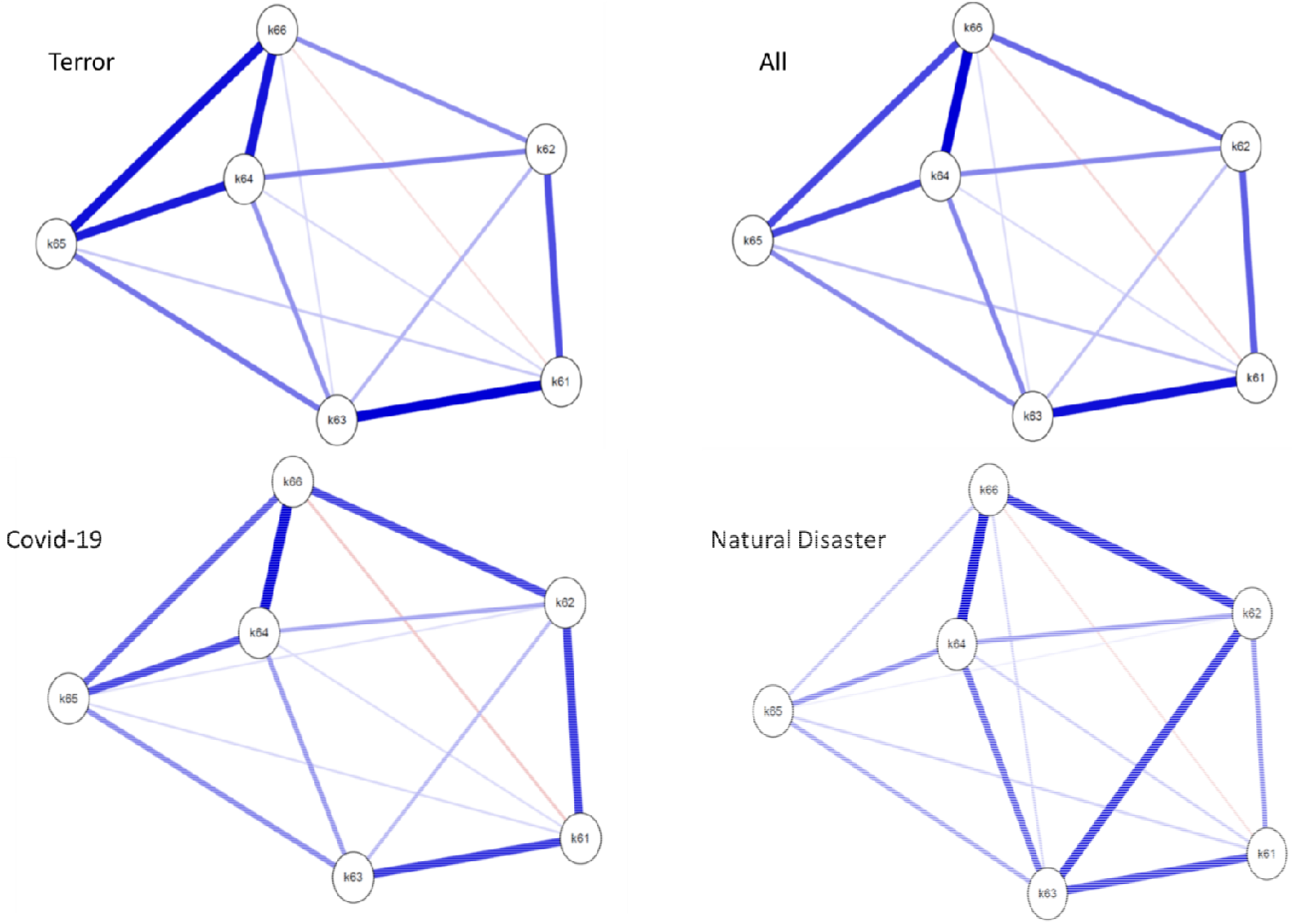
Networks of K-6 symptoms in three types of disaster samples using average spring layout. Nodes represent K-6 items, and edges Regularized partial correlations with LASSO penalty. Distances among nodes and thickness of edges relate to the size of their partial correlations. Blue edges indicate positive relations and Red edges indicate negative relationships. K61: nervous, K62: hopeless; K63: restless or fidgety, K64: so depressed that nothing could cheer you up; K65: that everything was an effort, K66: worthless

Using the total sample (all types of disasters) strong associations were found between the first item (“nervous”) and third item (“restless or fidgety”) and between item 4 (“depressed/no cheering up”) and both items 5 (“everything was an effort”) and 6 (“worthless”). However, visual inspection of the three separate networks shows inconsistent edges across the three subtype disasters with the only similarity being between the fourth (“depressed/no cheering up”) and sixth (“worthless”) item. In the Terror network, item 4 (“depressed/no cheering up”) was highly associated with both items 5 (“everything was an effort”) and 6 (“worthless”), and items 5 and 6 were also highly associated. Furthermore, in the Terror network, the first (“nervous”) and third (“restless or fidgety”) items associated highly compared to this association in the other networks which was strong but to a lower extent. In the Nature related disaster network, this triangle of items 4, 5, and 6 was substantially weaker, and only items 4 and 6 were strongly associated. In the COVID-19 and the Nature related disaster networks, item 2 (“hopeless”) was highly associated with item 6 (“worthless?”). In the natural disaster sample, item 2 (“hopeless”) was also highly associated with item 3 (restless or fidgety”). In the COVID-19 network, item 2 (“hopeless”) was also highly associated with the first item (“nervous”).

#### Network stability

To confirm this visual similarity of networks, we used Spearman correlations of edge-weights for all combinations of networks. Analysis shows that the accuracy of the edges was satisfactory. In addition, the results showed high accuracy of the centrality strength index with 85% of the edges that were nonzero had confidence intervals that did not contain zero. The stability coefficient for the strength centrality metric was above the suggested 0.5 cutoff for strong stability for all networks (0.75 CI 95% 0.67, 1.00). The correlation of the original strength centrality with the strength centrality in subsamples was high, suggesting the strength estimate can be considered stable in all three networks.

#### Network inference

The strength centrality estimates were similar across the three networks. Item 4 (“depressed/no cheering up”) was the node with the highest strength centrality in the total sample, in the Terror network as well as, in the COVID-19 network. In the Nature related disaster network, the most central item was item 3 (“restless or fidgety”) with 1.11 strength, and another item that had a similar strength centrality was item 4 (‘feel depressed/no cheering up’) with 1.09 centrality.

#### Network comparison between the three types of Disasters

Results from the network comparison test showed similar that global strength values per group were 2.83, 2.74 and 2.89 for Terror, Nature related disaster, and COVID-19, respectively (S statistics for each pair of samples ranged 0.05 to 0.15 and p value ranged .21 to .60). The network structure differed between the Terror and Nature related disaster networks (*M*=.26 *p*=.00), between the COVID-19 and Nature related disaster networks (*M*=.21 *p*=.00), as well as between Terror and the COVID-19 networks (*M*=.14 *p*=.00).

## Discussion

To our knowledge, this is the first investigation of the *K-6* network structure of psychological distress explored following three different types of disasters. Results suggest that K-6 psychological distress is disaster-specific, with network structure differing across three disasters types, reflecting different clinical manifestations of distress. While in the wake of a nature related disaster and the COVID-19 pandemic depression related and anxiety related items were interconnected, a terror attack resulted in more separated manifestations of anxiety and depression. Furthermore, centrality analysis showed that while “depressed/no cheering up” had the highest strength centrality in the Terror network and the COVID 19 network, for the Nature related disaster network, two items were most central: “depressed/no cheering up” and “restless or fidgety”. Networks were stable with high stability of the edges and good accuracy of the centrality index.

### The pattern of interactions between the K-6 symptoms in the combined sample

Examination of the symptoms network including participants of all disaster types revealed strong associations between item 4 (“depressed/no cheering up”) and both items 5 (“everything was an effort”) and 6 (“worthless”), all of which represent aspects of depression. Moreover, strong associations were found between item 1 (“nervous”) and item 3 (“restless or fidgety”), representing anxiety. In between the depression and anxiety items, the associations were weaker. This finding is in line with previous studies that identified two factors in the K-6, namely depression and anxiety ^8,9^. This pattern represents comorbidity between depression and anxiety. The observation that psychological distress as assessed by the K-6 is not a homogeneous construct is supported by studies showing that once comorbidity develops, the probability of recurrence of either disorder alone becomes low ^47^. Longitudinal network research on comorbid depression and anxiety also suggests that while bridging symptoms activate depression and anxiety and maintain the comorbidity, comorbidity can occur without overlapping bridging symptoms ^48^, similar to our study’s finding. This supports the argument that anxiety and depression may have overtime different expression, rather than comprising independent disorders ^47^.

### Network associations across different types of disasters

The symptoms of the K-6 had extensive connections with each other in all samples. The most consistent similarity between the networks was found to be the strong association between two depression items, namely Item 4 (“depressed/no cheering up”) and Item 6 (“worthless”). This suggests an overarching theme in the manifestation of distress following different disaster types. The significance of worthlessness as a symptom of distress was also confirmed in a network analysis of depression symptoms ^49^, indicating that it is closest to other symptoms and ‘bridges’ between nodes in a network. Feelings of inadequacy/worthlessness, depressed mood, and hopelessness also emerged as the most closely co-occurring and consistent symptoms among patients with remitted major depressive disorder ^50^. Finally, worthlessness was also found to be the most central symptom also for other stress-related disorders such as Complex PTSD ^43,51^.

However, symptom networks across types of disasters also showed significant differences in network structures. In the Nature related and COVID networks, worthlessness was strongly associated with both “depressed/no cheering up” and hopelessness. In the Terror network, on the other hand, “everything is an effort” was a core item strongly associated with “depressed/no cheering up” and worthlessness. It thus seems that while the Nature related and the COVID networks are characterized by depression-related elements that are associated with a negative self-concept and negative perception of the future, the Terror network was characterized by a depressive syndrome more behavioral and somatic in its expression, suggesting reduced energy or fatigue. The latter finding is supported by earlier study of the Charlie Hebdo terror shooting in France, which showed the rise in the probability of feeling depressed was 183%, with somatic expressions of depression being highly prevalent (e.g., requiring effort to live in 70%, feelings of restlessness in 90%) ^52^.

In the Terror network, the two anxiety items (nervous and restless/fidgety) were weakly connected to the depression items whereas, in the COVID-19 network and Nature related disaster network, anxiety and depression items were more strongly associated. This implies that the psychological reaction to COVID-19 and the Nature related disaster is well-captured by the construct of psychological distress, which amalgamates depressive and anxious symptoms. In the aftermath of terror, however, separate anxiety and depression syndromes seem to emerge. Person-made disasters such as terror attacks more frequently result in PTSD, a fear-related disorder, and in full-blown depression as comorbidity, compared to nature related disasters ^53^.

Associations between “everything is an effort” and both “depressed/no cheering up” and “worthlessness” were stronger in the COVID-19 network than in the Nature related network. We suggest that the unique stressors connected to COVID trigger the vulnerability for this depressive pattern. The detrimental impact of disasters is exacerbated if people perceive that trusted authorities had failed to react adequately, as has been reported for the COVID-19 pandemic. COVID-19 research showed that perceived institutional betrayal during the initial stages of the pandemic was the strongest predictor of negative affect in both Swiss and Israeli samples ^54^.

Furthermore, research on this pandemic has also identified social isolation as an important factor regarding mental health and wellbeing during this period of time. Although quarantine is common during infectious outbreaks ^55^, the COVID 19 pandemic was a rare example of widespread lockdown and social distancing ^55^. As worldwide cases of COVID-19 continue to increase, there is a risk that prolonged isolation and loneliness will intensify suicide and self-harm ^56^.

### Most central symptoms across networks

The centrality index, which reveals the highest level of clinical information, showed that “depressed/no cheering up” was the node with the highest strength centrality in the Terror network as well as in the COVID-19 network. Terror attacks and the varying stages of COVID-19 can breed a continuing sense of uncertainty. Also, both these events involve potential violations of interpersonal relationships and trust, particularly towards strangers, as it is the humans being the threat, not nature. A study of Chinese Hong Kong citizens during COVID showed that lack of perceived interpersonal trust was associated with probable depression ^57^. A sense of interpersonal isolation and uncertainty may be expected when an event originating from human activity undermines someone’s more benign viewpoint of their world ^29^. In order to reduce psychological distress, an intervention should aim to lower depressive affect, and to increase a sense of closeness and trust in others despite the necessary social distancing. This could be achieved, for example, by creating an atmosphere of togetherness against the threat, and social responsibility.

Contrarily, the results showed that in the Nature related disaster network two items were most central: “depressed/no cheering up” and “restless or fidgety”. This indicates the clinical import of both depression and anxiety. Compared to the other disaster types nature related disasters may particularly impact on the poorest communities most heavily ^23^ with post-disaster stressors (e.g., displacement to refugee camps, inability to get basic needs met, financial hardship, and difficulties rebuilding), accounting for more variance in psychological distress compared with the actual experiences of injury, loss, and threat resulting from direct exposure ^58–60^. These characteristics of natural disasters may explain the higher centrality of anxiety in the network.

Several limitations of this study should be taken into consideration. A potential bias in the present study is the use of self-report questionnaires. Second, the cross-sectional nature of networks in this study cannot infer causality ^61^ or predict bridging symptoms. Third, disaster samples were collected in different parts of the world and thus, cultural differences could provide an alternative explanation for differences found. Fourth, in the current study we used single disaster categorizations: Disasters however can also cascade so that a ‘natural’ event combines with human actions to provide a mass threat (e.g., in case of Fukushima, or when infectious diseases follow natural events ^23^. Future research is needed to further explore differences and similarities in psychological distress after multiple disasters.

Our study has important clinical implications. Trauma-focused interventions need to be augmented with specific efforts focused on depression and/or anxiety or psychological distress. Following nature related disasters interventions should focus on reducing both depressive and anxiety symptoms. However, after terror attacks or a novel pandemic, psychological distress may be addressed by reducing uncertainty and promoting trust and implementing interventions focusing on depressive affect and an individuals’ ability to savor those aspects of life untouched by the disasters.

## Data Availability

Data will be provided upon request from the last author 9Prof. Menachem Ben-Ezra)

