## Supplementary material for "Symptoms network analysis of serious mental illness: A cross disasters comparison": SM

Supplementary Materials

Yafit Levin^1^, Bachem Rahel^1^, Robin Goodwin^2^, Menachem Ben-Ezra^3^

^1^ University of Zurich, Switzerland

^2^ Department of Psychology, University of Warwick, Coventry, UK

^3^ School of Social Work, Ariel University, Ariel, Israel

Correspondance : Dr. Yafit Levin.

**Terror** **samples from France, the UK, and the US**

**France.** The sample comprised of two data collections referring to two terror attacks that took place in 2015. The Charlie Hebdo shootings took place from 7-9th January 2015 and associated suicide bombings took place on 13^th^ of November 2015. A major survey company was employed to collect data during seven days, four weeks after each attack, drawing on an established internet panel of almost half a million participants across France. For each event, samples were selected from an existing panel using random stratified sampling methods, using weights for key demographic elements (age, sex) that were compared with French census information to create a reliable approximation of a representative sample. All respondents were aged over 18.

During the week of 8th February 2015, 6059 survey panel members were sent a web-link of which 2421 clicked through to the survey. 1981 (82%) of these passed a validation question and responded fully. A month after the November mass attacks 2015 (week of December 13, 2015) 2612 panel members were sent the web link, of whom 1878 passed a validation question and participated fully (response rate 72%). There were no significant differences between the two samples, which were summed to 3456 participants.

**UK.** The sample comprised of a data collection referring to a terror attack that took place on May 22, 2017, in which a suicide bomber exploded during a concert in the Manchester Arena. An online sample targeting the UK population was conducted during 25-29 May, 2017. The inclusion criteria were age of 18 and above and fluent in English. We used a UK based survey company (surveygoo) to deploy the survey among the UK population. Each participant received an invitation to participate in the study and signed an electronic informed consent. Out of 1710 invitations sent, 1504 answered the questionnaire, after omission of participants with missing data, a final sample of 1399 participants who have fully answered the survey (response rate = 81.8%). Further information on demographic variables can be found in (Ben-Ezra, Hamama-Raz, & Mahat-Shamir, 2017).

**US**. The sample comprised of a data collection referring to a terror attack that took place on June 12, 2016, a combined terrorist and hate crime attack targeted a nightclub in Orlando, Florida. Three weeks after the mass shooting, we conducted a study with a state census representative sample (meeting Florida census proportions for age and gender) taken from an online panel. From 1350 invitations sent, 987 respondents answered the survey fully and their data were eligible for analyses (response rate = 73.1%). The study was approved by the ethics committee in the Faculty of Social Science and Humanities at Ariel University. More information on demographics can be found in (Ben-Ezra et al., 2017).
